# Identifying Training Needs of Healthcare Providers to Implement Caries Risk Assessment

**DOI:** 10.1101/2025.05.29.25328525

**Authors:** Olubukola O. Olatosi, Robert J. Schroth, Daniella DeMaré, Maria Manigque, Betty-Anne Mittermuller, Jeanette Edwards, Katherine Yerex, Peter Wong, Josée Lavoie, Julianne Sanguins, Prashen Chelikani, Alexandra Nicolae, Jesse Lamoureux, Rhonda Campbell, Mary Bertone, Maryam Amin, Working Together for early childhood oral health study team.

**Affiliations:** Department of Preventive Dental Science, Dr. Gerald Niznick College of Dentistry, Rady Faculty of Health Sciences, University of Manitoba, Winnipeg, Manitoba, Canada; Department of Oral Biology, Dr. Gerald Niznick College of Dentistry, Rady Faculty of Health Sciences, University of Manitoba, Winnipeg, Manitoba, Canada; Children’s Hospital Research Institute of Manitoba, Winnipeg, Manitoba, Canada; Department of Pediatrics and Child Health, Max Rady College of Medicine, Rady Faculty of Health Sciences, University of Manitoba, Winnipeg, Manitoba, Canada; Shared Health, Winnipeg, Canada; School of Dental Hygiene, Dr. Gerald Niznick College of Dentistry, Rady Faculty of Health Sciences, University of Manitoba, Winnipeg, Canada; Department of Pediatrics, University of Toronto, Toronto, Ontario, Canada; Department of Community Health Sciences, Max Rady College of Medicine, Rady Faculty of Health Sciences, University of Manitoba, Winnipeg, Manitoba, Canada; Ongomiizwin Research – Indigenous Institute of Health and Healing, Rady Faculty of Health Sciences, University of Manitoba, Winnipeg, Manitoba, Canada; Toronto Public Health, Toronto, Ontario, Canada; Interlake-Eastern Regional Health Authority, Pine Falls, Manitoba, Canada; College of Nursing, Rady Faculty of Health Sciences, University of Manitoba, Winnipeg, Manitoba, Canada; School of Dentistry, University of Alberta, Edmonton, Alberta, Canada

**Keywords:** Indigenous oral health, early childhood caries, caries risk assessment, health care providers, pediatric primary care

## Abstract

**Background:** Early childhood caries remains a pressing concern among Indigenous children in Canada, driven by systemic inequities, limited access to care, and fragmented service delivery. Integrating caries risk assessment (CRA) into primary care presents an opportunity to improve early detection and prevention. This study explored the training needs and preferred delivery methods of non-dental primary care providers (NDPCPs) to support CRA implementation in Indigenous pediatric settings.

**Methods:** This qualitative exploratory study involved 50 NDPCPs serving First Nations and Métis children under six years of age across 10 communities in Manitoba. Data were collected between April 2023 and February 2025 through eight focus groups and 12 key informant interviews, followed by brief individual interviews to assess preferred training modalities. Transcripts were analyzed using thematic analysis to identify key training needs and preferences.

**Results:** Participants included physicians, nurse practitioners, public health nurses, physician assistants, dietitians, and child development workers. Four core training areas were identified: dental caries screening, CRA tool usage, fluoride varnish application, and documentation/referral processes. Despite recognizing the CRA tool’s value and ease of use, participants reported limited formal training in preventive oral health and stressed the need for hands-on, culturally appropriate instruction. Preferred training modalities varied by geography: urban providers favored blended in-person and online approaches, while rural providers preferred online formats due to travel constraints. Overall, in-person and interactive training was most preferred.

**Conclusion:** NDPCPs require structured, context-specific training to effectively integrate CRA into routine care. A hybrid training model combining online modules with locally delivered, hands-on learning may best address geographic and resource-based disparities. Training content should be simple, skill-focused, and culturally responsive to support NDPCPs in delivering equitable oral healthcare to Indigenous children.

## Introduction

Dental caries, commonly known as tooth decay, remains a pressing public health issue that disproportionately affects Indigenous communities in Canada (1, 2). Despite advancements in preventive care and education, Indigenous populations continue to experience higher rates of dental caries compared to the general population. Contributing factors include limited access to oral health services, socio-economic challenges, cultural barriers, provision of culturally inappropriate services and the lasting impact of colonization (3–5). For those residing in rural and remote areas, additional barriers such as a shortage of dental professionals, geographic remoteness, financial constraints, travel difficulties, inadequate infrastructure, and limited dental insurance coverage further exacerbate oral health disparities (6, 7). Moreover, the fragmentation of healthcare services and the separation of dental and medical care have intensified the burden of oral disease among Indigenous populations (8, 9). Addressing this inequity requires innovative strategies that integrate oral healthcare into existing primary care frameworks.

Non-dental primary care providers (NDPCPs) play a crucial role in delivering comprehensive health services within Indigenous communities. As frontline healthcare professionals, they are well-positioned to identify and address oral health risks (10, 11). Collaborative efforts between medical and dental professionals can enhance service delivery, ensuring that underserved populations receive timely oral health assessments and interventions (12). Several professional organizations advocate for medical providers to conduct oral health assessments for children as young as six months and address early childhood caries (ECC) risk factors (13–15). Additionally, the US Preventive Services Task Force (USPSTF) recommends that primary care clinicians apply fluoride varnish to the primary teeth of all infants and children starting at the age of primary tooth eruption (Grade B) (16).

The Canadian Caries Risk Assessment (CRA) tool for children below age 6 years is a standardized tool designed to assess an individual’s risk for developing dental caries and guide preventive strategies (11). However, successful implementation of this tool within Indigenous communities requires targeted training for NDPCPs to ensure effective utilization while maintaining cultural appropriateness.

This paper explores (1) the training needs of NDPCPs and (2) their preferred training delivery method for the implementation of the CRA tool in Indigenous communities. By identifying knowledge gaps, skill requirements, and cultural safety needed, this research aims to provide actionable recommendations to strengthen the capacity of primary care providers in addressing oral health disparities. It further emphasizes the importance of a collaborative, community-centered approach to ensure that training programs are aligned with the unique needs, preferences, and values of Indigenous populations.

## Methods

### Ethical Approval

This study received ethics approval from the University of Manitoba Health Research Ethics Board (HREB) under approval number HS25866 (H2023:050). The study is part of a broader research initiative funded by the Canadian Institutes of Health Research (CIHR) in partnership with the First Nations Health and Social Secretariat of Manitoba (FNHSSM) and the Manitoba Métis Federation (MMF), with additional ethics approvals HS24621 (H2021:043) and HS20926.

### Study Design and Setting

This qualitative exploratory study aimed to identify the training needs and preferred training delivery methods for NDPCPs working with First Nations and Métis children in Manitoba to implement and integrate the Canadian CRA tool (Fig. 1) into primary care. The study builds upon previous research that identified barriers to implementing the CRA tool for preschool-aged children (17).

**Fig 1:**
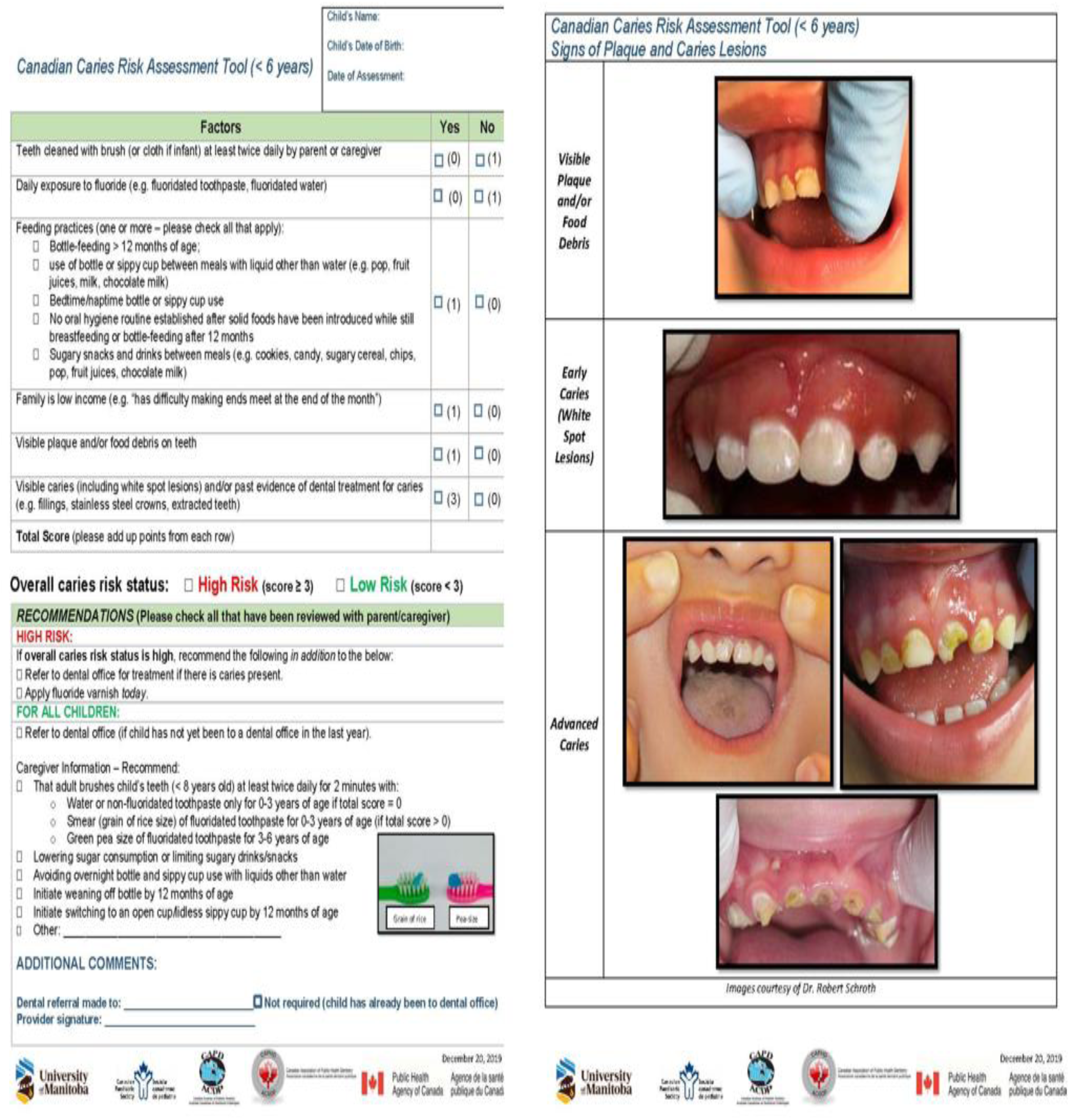
Canadian Caries risk Assessment Tool for preschoolers

### Participant Recruitment and Selection

Participants were purposefully recruited based on predefined eligibility criteria. Inclusion criteria required that participants be NDPCPs serving First Nations and Métis children under six years old. Recruitment took place across 10 Indigenous communities in Manitoba, and details of the recruitment process have been previously reported (17).

### Data Collection

Between April 2023 and September 2024, 50 NDPCPs participated in the study through eight focus groups and 12 in-depth key informant interviews. A semi-structured interview guide, informed by a literature review and previous research, was iteratively refined by an interdisciplinary team specializing in early childhood oral health, health promotion, community development, and Indigenous health. Focus group sessions, conducted at community health centers, lasted between 45 to 75 minutes, while key informant interviews lasted 15 to 30 minutes. Sessions were audio-recorded and supplemented with field notes. A follow-up interview phase, conducted between December 2024 and February 2025, sought to determine participants’ preferred training delivery methods. Participants were asked to indicate their preferred format among the following three training modalities (18):

1. Online Training – Internet-based learning, including slide presentations, written modules, online videos, or webinars.
2. In-Person Training – Face-to-face lectures, often including Q&A sessions.
3. Either 1 or 2 inclusive of Interactive Training – A supplementary component involving hands-on demonstrations (e.g., patient positioning for oral screenings or fluoride varnish application).

The follow-up interviews averaged five minutes, and all interviews were transcribed verbatim.

### Data Analysis

Data analysis was conducted concurrently with data collection, employing both inductive and deductive thematic approaches. We used the constant comparative method to iteratively refine emerging themes and concepts. Open coding was done by analyzing transcript line by line, with codes assigned to capture key meanings. Thematic development was done by grouping related codes to form themes, ensuring a structured interpretation of the data.

### Rigor and Trustworthiness

To ensure methodological rigor, the study adhered to qualitative research standards emphasizing dependability, confirmability, credibility, and transferability (19). Measures such as audio recordings, verbatim transcriptions, and participant verification enhanced data reliability. Detailed methodological descriptions and direct quotes were included to support transferability (20).

## Results

Interviews were conducted between April 2023 and February 2025, with a total of 50 NDPCPs participating in the baseline interviews. The participants represented a diverse range of healthcare professionals, including physicians (n=8), public health nurses (n=31), nurse practitioners (n=4), physician assistants (n=2), dietitian (n=1), and child development workers (n=4). Through thematic analysis, four key themes emerged regarding the training needs of participants: screening for caries, CRA screening tool, fluoride varnish application, and documentation and referral pathways.

At follow-up, 28 NDPCPs participated, with 60% practicing in urban centers. Geographic location was considered a potential factor influencing preferred training delivery methods. Most participants (64%) preferred an in-person plus interactive training approach, followed by online plus interactive training (60%). The least preferred method among participants was the online-only format. Notably, among NDPCPs in rural centers, 71% preferred online-only training, highlighting the impact of geographic constraints on training preferences (Fig. 2).

**Fig 2:**
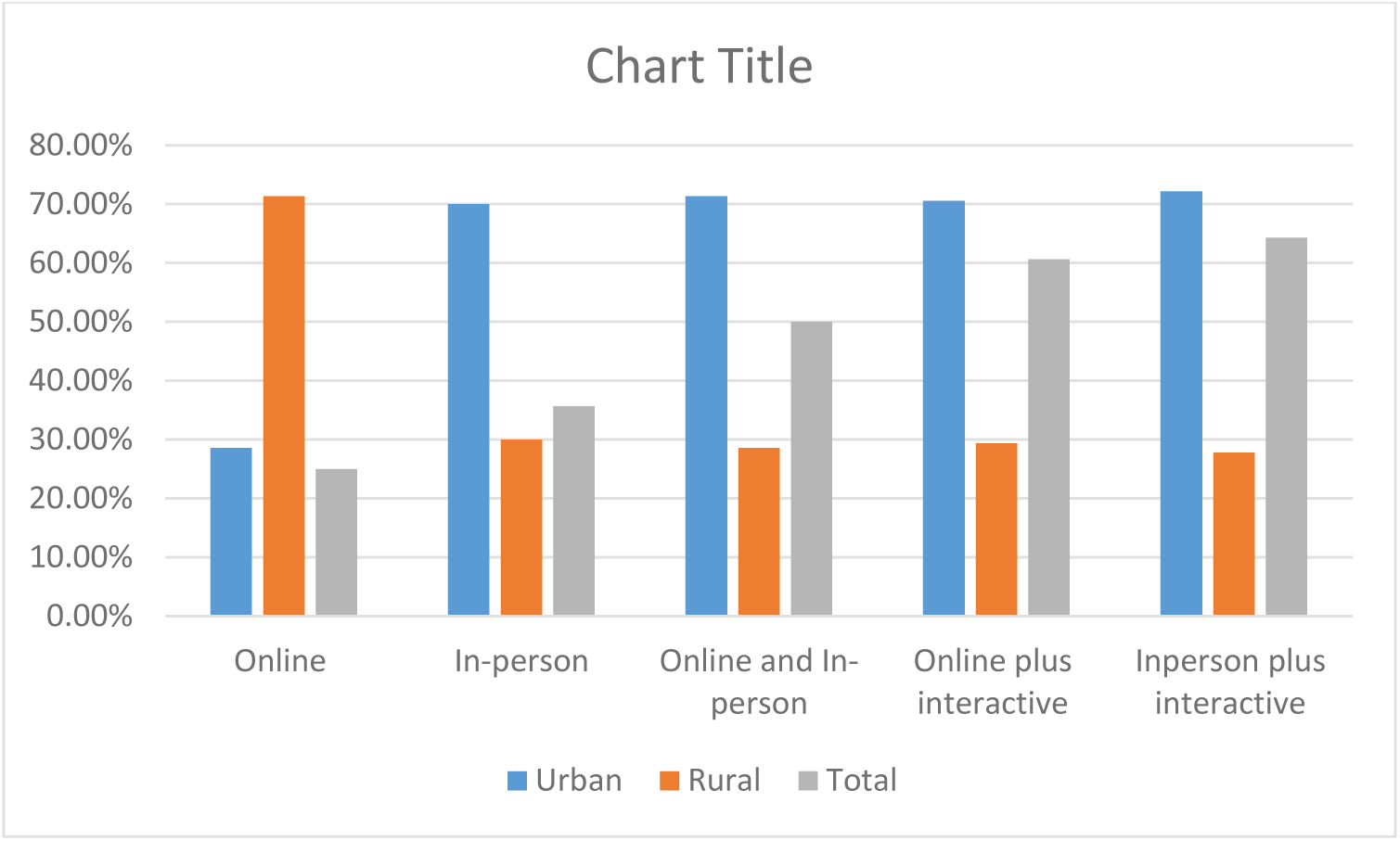
Geographic Classification and Preferred CRA Training Delivery Method

### Screening for Dental Caries

Participants emphasized the need for training on identifying and screening for dental caries. They highlighted the importance of understanding the different stages of caries progression, with some suggesting that visual aids such as pictures or opportunities to observe and shadow dental providers would be beneficial. Additionally, they expressed interest in training on the causes and prevention of dental caries. Given time constraints, they recommended that the training be kept simple to facilitate quick identification and screening.

> *"If you can get … experienced dental colleague to do very quick [training] because people don’t really have time for anything that takes too long. And done in such a way to improve comfort of primary care providers when dealing with oral health. So we need to keep it super and drill it down to the basics”* (Nurse Practitioner, 2).

One family physician, in particular, noted a lack of specific training in pediatric dental exams and expressed a need for additional education on the subject:

> *"As a family doc, we don’t have any information specific about the kid at that age about how to do the exam or what we are looking for. If we can have actual training or something like that will be beneficial for sure."* (Physician, 1)

Some participants stressed the importance of understanding not just how to screen for caries but also what interventions dentists provide and the potential consequences of untreated childhood dental caries:

> *"Getting some additional training on recognition of pediatric dental caries, some education on what the dentists do, what they offer and what kind of interventions they do, and some more education on the consequences of childhood dental caries just for education purposes for the parents would be helpful."* (Physician, 2)

Others suggested that training should be tailored to varying levels of confidence and comfort among healthcare providers. A mix of visual materials and hands-on demonstrations could be effective in ensuring competency:

> *"Everybody’s practice is different and everybody’s confident and comfort level is different. So I think even a visual like these [referring to the CRA tool] are great. So maybe more in detail would be helpful."* (Nurse Practitioner, 1)

Finally, participants underscored the importance of making training concise and accessible, with a focus on building confidence among primary care providers in addressing oral health:

> *"I think really need to be able to observe or shadow the dental hygienist or dentist when they screen preschoolers, that would be a good training to see the actual team person do it."* (Nurse, 1)

### Caries Risk Assessment (CRA) Tool

Participants generally found the Caries Risk Assessment (CRA) tool easy to use and well-structured. One nurse described it as straightforward:

> *"I would say a quick review, right? Like, this is very nicely laid out. I feel this is very straightforward"* (Nurse, 2).

However, while the tool itself was considered user-friendly, participants highlighted the need for initial training to familiarize themselves with its content, particularly regarding anticipatory guidance and answering related questions from parents. As one nurse explained:

> *"Just the confidence, right? Like getting familiar with the tool, getting comfortable using it, and then the varnish…remembering to do it as part of our regular assessment"* (Nurse, 3).

Some participants sought clarification on the tool’s practical application, including its frequency of use and expected outcomes:

> *"How do we navigate through this form? How often do you want us to fill this form and what is the outcome of this form? How do we appreciate there is outcome of this work"* (Physician, 3).

Sustaining and maintaining training for new staff was also identified as a key factor in successful implementation. Participants emphasized the importance of ongoing education, particularly in ensuring cultural competence when using the tool:

> *"We’d have to get a good overview education for whoever’s providing it. And then keeping on top of that, for all the new staff that come in as part of the implementation process. How to use the tool and how to talk to them about it? So it’s about making sure that they’re just making an informed decision, not being judgmental"* (Nurse Practitioner, 3).

Finally, participants acknowledged that proper training over time builds confidence in using the CRA tool effectively:

> *"Just like with the appropriate training, I think, you know, having that training builds the confidence and with time, over time"* (Dietitian).

Overall, while the CRA tool was seen as an accessible and valuable resource, participants emphasized the importance of initial and ongoing training to enhance confidence, ensure accurate risk assessment, and maintain cultural sensitivity in its application.

### Fluoride Varnish Application

Participants discussed their knowledge of fluoride and fluoride varnish application as a crucial skill necessary for implementing the CRA tool. While many recognized the need for training in fluoride varnish application, some questioned whether it fell within their job responsibilities. However, they expressed willingness to undergo training if it were clearly outlined in their job descriptions and supported by management. As one participant stated:

> *"Like you said, it’s within our scope, but we’re nurses and nowhere in nursing school that I learned to apply varnish and even for public health. So we would need training, even if it’s very like simple. And told by our up above that it is what we are doing"* (Nurse, 4).

Another important concern identified by participants was the behavioral management of children during fluoride varnish application. Some expressed the need for strategies to ensure children’s cooperation, as it can be challenging to keep their mouths open long enough for application. One nurse highlighted this challenge:

> *"And regarding the fluoride [varnish] about applying it, I would definitely say we would want some training and maybe tips on how to get a kid to keep their mouth open long enough for me to put fluoride. I would like some tips and tricks on how you guys manage that"* (Nurse, 5).

Beyond the technical aspect of fluoride varnish application, participants also noted gaps in their knowledge regarding the benefits of fluoride and how to effectively communicate these benefits to parents. Some emphasized the importance of having clear post-application instructions available in the form of flyers or leaflets for parents and caregivers. One participant expressed a need for more detailed guidance:

> *"If we were to apply fluoride, I guess I would need some more information about how to do that. If they’re [children] supposed to spit it out, swallow it and leave it on like I don’t know anything about what’s supposed to be done with that"* (Nurse, 6).

Additionally, some participants believed that training and implementation should be driven by higher-level management decisions. They indicated that while they are involved in patient education, direct application of fluoride varnish would require structured training and support from leadership:

> *"That would definitely be coming from much higher up for that part, because we do the education pieces but yeah, applying the fluoride varnish, I would want training if it was implemented through management."* (Nurse, 7)

Overall, the discussion highlighted the need for structured training in fluoride varnish application, behavioral management strategies for children, improved knowledge on the benefits of fluoride, and clear communication tools for parents. Participants emphasized the importance of management support in successfully integrating fluoride varnish application into their practice.

### Documentation and Referral

Participants discussed the challenges and training needs associated with documentation and referral for dental care, particularly for their patients who are predominantly from low socioeconomic backgrounds. They emphasized the need for structured training on how to document referrals effectively and clear information on available and affordable treatment centers to ensure that patients receive appropriate dental care.

> *"Just knowing which centers are going to be available for patients, particularly of low socioeconomic class, where they can go and get affordable treatment, just that sort of information will be useful."* (Physician, 4)

Many participants expressed uncertainty about the referral process, including where to send patients and the affordability of services. They highlighted the need for training on referral pathways, eligibility criteria, and financial assistance programs:

> *"Oh, refer to dental office for treatment. You know what? That’s another thing, sometimes I’m like who are we referring them to, where are they going? Is it free? Is there low budget friendly, low income ….That’s always the issue, I never really know"* (Nurse, 8).

Participants also pointed out challenges in ensuring timely follow-up after an initial screening, often resulting in delays in care. Training in effective coordination with dental professionals and streamlining referrals was identified as a key need:

> *"We need better access to the professional screeners or to the appropriate dentist, or dental hygienist because we’ve run into barriers where we could screen but then the follow-up or to refer them to another professional could take a little longer"* (Nurse, 1).

Ensuring a positive experience for referred families was another concern. Participants emphasized the need for training on how to identify and refer patients to trusted and culturally safe dental providers who understand the unique challenges faced by low-income families and provide a welcoming, nonjudgmental environment:

> *“I would want to make sure that this family is going to have a good experience and without knowing certain dentists, how do I pick one over another? If there is a list of dentists who were aware of this and who praise families for coming in, instead of why did you wait so long? I think that would be helpful"* (Nurse, 9).

Overall, participants highlighted the need for structured training on documentation and referral processes as part of the implementation of the CRA tool. This includes knowledge of available dental resources, referral pathways, and strategies to ensure timely and effective follow-up. Addressing these training needs would enhance the efficiency of the referral process and improve access to dental care for vulnerable populations.

### Preferred delivery method for CRA training

### Online-Only Training

Online-only training was the least preferred method, selected by 25% of participants. However, among those who chose this method, 71% were located in rural areas of Manitoba. Distance and accessibility were key considerations for these participants:

> *“Online because of where we live. For fluoride varnish application, it depends if they can drive out to us. If we have to drive to them, I would prefer all of the training to be online”* (Child Development Worker, 4).

#### In-Person Training

Thirty-six percent of participants preferred in-person training, citing benefits such as a more engaging learning environment and greater interaction:

> *“Personally, in person is a better learning environment because the information is right in front of your face”* (Nurse, 12).

> *“I’m more of an interactive learner, so I would prefer training to be in-person. For fluoride varnish, I would prefer that to be demonstrated in person”* (Physician, 5).

Some participants suggested structuring in-person training around their work schedules:

> *“In our clinic, it’s very busy, but it’s best to have this training in person. Nurses have a protected lunchtime, but I think this should be done over lunch and stagger the training sessions. I would suggest talking to the clinic manager”* (Nurse, 13).

#### Online and In-Person Training

Half of the participants preferred a blended approach, integrating both online and in-person training. Among these, 70% were from urban centers. Participants recommended using online training for didactic instruction and content review, while in-person sessions could focus on hands-on experience and case discussions:

> *“I think online would be great to review the information, and in-person would be good for hands-on experience mixed with case studies”* (Nurse, 11).

> *“Online didactic training should be one hour max. Practical training should be in person”* (Physician, 6).

#### In-Person plus Interactive Training

This was the most preferred training method, chosen by 64% of participants. Many emphasized the importance of hands-on learning for skill development:

> *“I’m more of an interactive learner, so I would prefer training to be in-person. For fluoride varnish, I would prefer that to be demonstrated in person”* (Physician, 5).

> *“A lot of people need hands-on training. A lot of people are visual learners and need that in-person training”* (Nurse Practitioner, 4).

> *“Hands-on would allow me to get a feel for what’s expected of me. For fluoride varnish, I would like in-person demonstrations with practice”* (Nurse, 8).

## Discussion

This study explored the training needs and preferred delivery methods for implementing CRA tool among NDPCPs in Indigenous pediatric primary care settings. Key findings from this present study include strong endorsement of the CRA tool’s usability and value, significant training gaps in preventive oral health skills, a preference for culturally relevant and hands-on learning, and the need for a hybrid training model that accommodates geographic and resource differences. These findings will be useful in informing the next steps of integrating CRA into primary care settings in Manitoba.

In the current study, participants described the CRA tool as beneficial for children, easy to use, and visually accessible due to its clear instructions and use of pictures. This is encouraging feedback as our CRA tool’s design was shaped by feedback from 63 NDPCPs and stakeholders. Key suggestions from that earlier study emphasized the importance of a tool that is quick to complete, easy to score, seamlessly integrated into clinical workflows, and inclusive of anticipatory guidance for parents and caregivers (21).

While NDPCPs in this study had positive feedback on the CRA tool, they also identified significant training gaps that need to be addressed for CRA to become part of their daily practices. These included limited knowledge and skills related to dental caries screening, the use of the CRA tool itself, fluoride varnish application, and documentation and referral procedures. These findings are consistent with earlier studies showing that NDPCPs often lack sufficient training in preventive oral health services (POHS), which restricts integration into routine care (9, 22). Participants in this study specifically noted the absence of formal training in POHS as a barrier that undermines their confidence and ability to conduct CRA effectively. Evidence suggests that incorporating oral health into the training curricula of NDPCPs can enhance providers’ confidence in delivering preventive oral health services, including anticipatory guidance, caries screening, and oral health risk assessments (23). Moreover, inter-professional collaboration and structured training programs have been shown to enhance the role of NDPCPs in oral health promotion (24–26).

Importantly, many participants noted that their willingness to participate in training depended on support from clinic leadership and alignment with their professional responsibilities. Leadership buy-in can support the necessary support, resources, authority and commitment to sustain change in practice (27). Also shared vision between management and staff is essential for managing practice change (28). A recurring concern was managing children’s behavior during fluoride varnish application, underscoring the need for training in child-centered communication strategies. Participants also called for clear guidance for parents’ post-application, including accessible educational materials, this aligns with previous studies (18, 29). Identifying these perceived training needs is a crucial first step towards implementing the CRA tool effectively.

Providers highlighted the need for culturally grounded strategies, practical demonstrations, and strong leadership support. These findings echo previous studies emphasizing that collaborative, culturally responsive approaches are critical for the successful implementation of oral health interventions in Indigenous contexts (30, 31). Sustained and tailored training, particularly for new staff members, was considered essential to ensure continuity of care.

Training needs extended beyond clinical competencies to include navigating local dental systems, understanding eligibility for publicly funded programs, and clarifying referral processes. These concerns reflect systemic barriers frequently cited in the literature, including poor access to dental providers and ambiguous referral pathways (32, 33).

The present study also identified distinct preferences for training delivery methods among the NDPCPs. Training delivery preferences varied based on geography and resource availability. Overall, participants preferred in-person and interactive training methods, citing the value of hands-on learning, real-time interaction, and opportunities to practice new skills. NDPCPs in urban areas favored blended training models, combining online theory with in-person skill development an approach supported by earlier research (34).

In contrast, rural providers leaned toward online-only formats due to travel and staffing constraints. Interestingly, these preferences diverge from trends in broader healthcare training. A study by Sams et al, found that online training was the most predominant method used in 32 U.S. States to educate NDPCPs in oral health, with blended approaches being the least utilized (18).

Online programs such as *Smiles for Life* (SFL) and *Protecting All Children’s Teeth* (PACT) are widely endorsed for their accessibility and flexibility. However, participants in our study emphasized the greater value of experiential learning, hands-on learning over passive didactic methods.

Interactive modalities such as role-playing, case discussions, and skill demonstrations have been shown to significantly improve provider confidence and performance (35, 36). While online training was the least preferred option overall, it was endorsed by rural participants for its flexibility and accessibility. The convenience of asynchronous, self-paced online learning is particularly beneficial in remote areas where staffing and travel barriers are more pronounced (37).

The growing popularity of online learning, accelerated by the COVID-19 pandemic, has ushered in new technological innovations, including simulations, digital teaching aids, and virtual learning platforms (38, 39). A recent systematic review concluded that blended or online training approaches can achieve comparable, if not superior, outcomes in clinical skills development compared to traditional methods (40).

Drawing on insights from this study, we recommend a geographically sensitive and needs-based approach to training NDPCPs for effective implementation of the Canadian CRA tool. A hybrid training model that combines established online modules such as SFL with locally accessible, in-person mentorship or practical demonstrations may offer the most feasible and impactful solution. This blended approach addresses both the flexibility required by providers in remote areas and the hands-on learning preferences expressed by many participants. To support efficient and sustainable integration of CRA into routine practice, training content should remain simple and focused. Core topics should include (Fig 3):

- An overview of ECC and prevention strategies
- Purpose and application of the CRA tool
- Child oral health screening and caries detection techniques
- Fluoride varnish application
- Communication with families and anticipatory guidance
- Guidance on establishing a dental home and navigating referral pathways

**Fig. 3.**
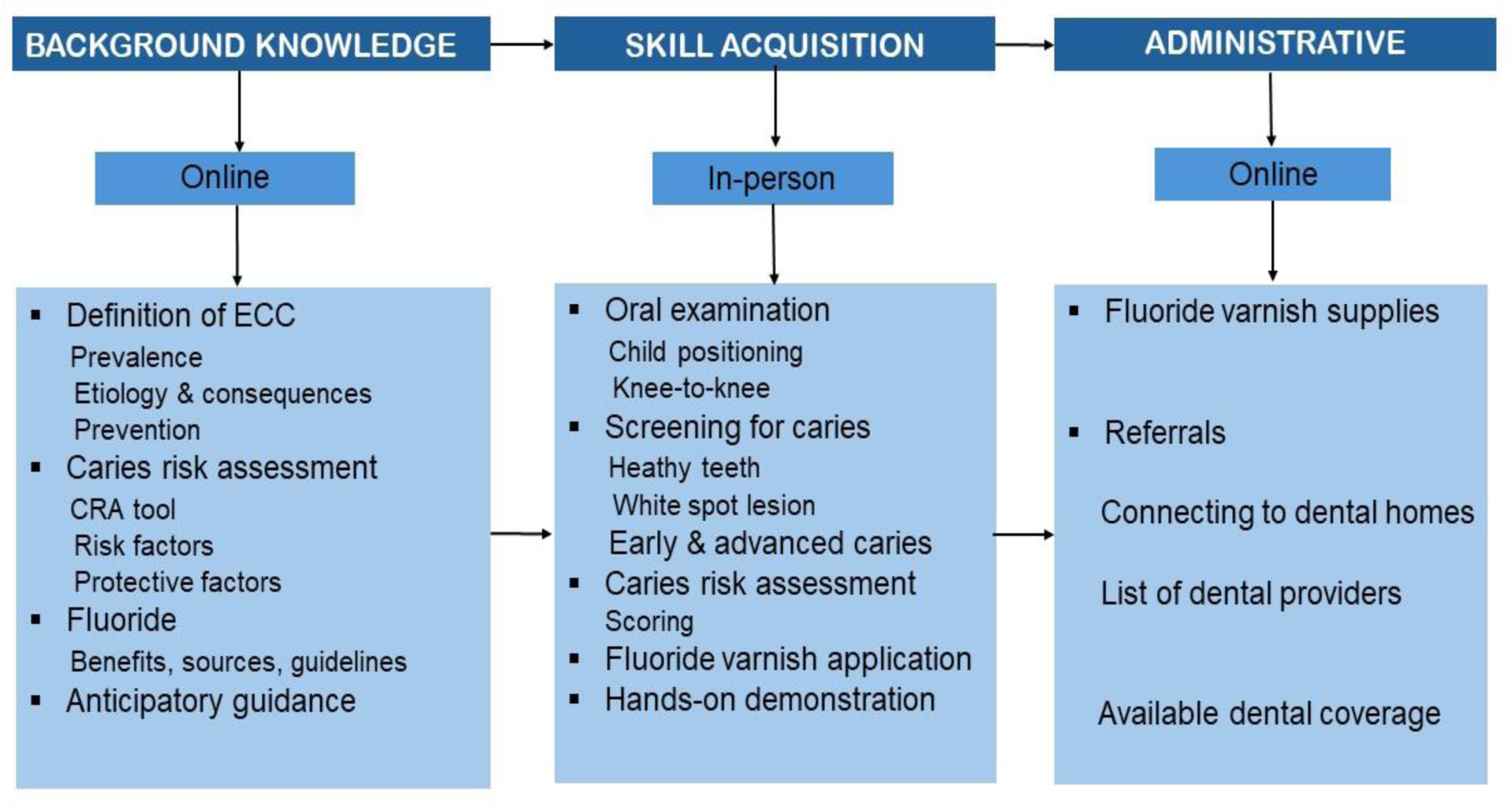
CRA Training Model

These targeted areas reflect both the learning needs identified by NDPCPs and broader best practices in preventive oral health care. Streamlined, practical training tailored to real-world clinical workflows can empower providers to confidently deliver oral health services and improve outcomes for Indigenous children.

### Strengths and Limitations

A strength of this study is its focus on the voices of NDPCPs working directly with Indigenous children and families. The inclusion of various professional roles and settings provides a rich, practice-based understanding of training needs. However, the study is limited by its focus on one province, which may limit generalizability. Additionally, while the logic model was developed with participant input, future work is needed to test its effectiveness in guiding CRA training implementation and evaluating outcomes.

## Conclusion

This study provides critical insights into the training needs and delivery preferences of NDPCPs for integrating the CRA tool into Indigenous pediatric primary care. Despite recognizing the tool’s value, providers expressed low confidence in performing preventive oral health tasks, pointing to the need for structured, context-specific training. A hybrid approach that combines online flexibility with in-person, hands-on learning tailored to both urban and rural settings is essential to build provider competence and confidence. Training must be simple, culturally grounded, and aligned with real-world workflows to support the successful integration of CRA into routine care. Empowering NDPCPs through targeted training and leadership support is a foundational step toward promoting oral health equity for Indigenous children across Canada.

## Author Contributions

O. O.O and R.J.S contributed to conception, design, data analysis interpretation, drafted and critically revised manuscript; D. D, B. M and M. M contributed to data acquisition, interpretation and critically revised manuscript. J. E, M. A, J. L, and J. S contributed to interpretation of results and critically revised manuscript; A. N, P. C and P. W critically revised manuscript; J. L contributed to data acquisition and critically revised manuscript; M. B and K. Y contributed to conception and critically revised manuscript; R. C contributed to data interpretation acquisition and interpretation. All authors gave final approval and agree to be accountable for all aspects of the work.

## Acknowledgments

The authors wish to thank all the NDPCPs who participated in this study.

## Data Availability Statement

The datasets generated and analyzed during the current study are available from the corresponding author on reasonable request. Data is available at Children’s Hospital Research Institute of Manitoba, University of Manitoba shared drive and will be shared on reasonable request.

